# Predicting the public health impact of bivalent vaccines and nirmatrelvir-ritonavir against COVID-19

**DOI:** 10.1101/2023.05.18.23289533

**Authors:** Hailey J. Park, Sophia T. Tan, Tomás M. León, Seema Jain, Robert Schechter, Nathan C. Lo

## Abstract

**Background:** Uptake of COVID-19 bivalent vaccines and oral medication nirmatrelvir-ritonavir (Paxlovid) has remained low across the United States. Assessing the public health impact of increasing uptake of these interventions in key risk groups can guide further public health resources and policy.

**Methods:** This modeling study used person-level data from the California Department of Public Health on COVID-19 cases, hospitalizations, deaths, and vaccine administration from July 23, 2022 to January 23, 2023. We modeled the impact of additional uptake of bivalent COVID-19 vaccines and nirmatrelvir-ritonavir during acute illness in different risk groups defined by age (50+, 65+, 75+ years) and vaccination status (everyone, primary series only, previously vaccinated). We predicted the number of averted COVID-19 cases, hospitalizations, and deaths and number needed to treat (NNT).

**Results:** For both bivalent vaccines and nirmatrelvir-ritonavir, the most efficient strategy (based on NNT) for averting severe COVID-19 was targeting the 75+ years group. We predicted that perfect coverage of bivalent boosters in the 75+ years group would avert 3,920 hospitalizations (95%UI: 2,491-4,882; 7.8% total averted; NNT 387) and 1,074 deaths (95%UI: 774-1,355; 16.2% total averted; NNT 1,410). Perfect uptake of nirmatrelvir-ritonavir in the 75+ years group would avert 5,644 hospitalizations (95%UI: 3,947-6,826; 11.2% total averted; NNT 11) and 1,669 deaths (95%UI: 1,053-2,038; 25.2% total averted; NNT 35).

**Conclusions:** These findings suggest prioritizing uptake of bivalent boosters and nirmatrelvir-ritonavir among the oldest age groups would be efficient and have substantial public health impact in reducing the burden of severe COVID-19, but would not address the entire burden of severe COVID-19.

## Introduction

The COVID-19 pandemic, caused by severe acute respiratory syndrome coronavirus 2 (SARS-CoV-2), continues to be a public health problem in the United States^1,2^. The epidemiologic landscape of COVID-19 in the United States has changed over the pandemic and is now characterized by a population with widespread vaccination with monovalent COVID-19 vaccines (including 69% fully vaccinated, and 38% with monovalent booster doses as of January 2023), high prevalence of prior infection, and emergence of increasingly infectious SARS-CoV-2 variants such as Omicron sub-variants^3,4,5^. As social distancing and public health measures are relaxed, a key public health question is understanding the impact of increasing uptake of additional vaccination and oral medications to further mitigate hospitalizations and deaths from COVID-19. Two key medical interventions against severe COVID-19 are bivalent vaccines and use of oral antiviral medications during COVID-19 illness, most commonly nirmatrelvir/ritonavir (Paxlovid)^6,7,8,9^; however, uptake of these interventions has been low^5,10^.

COVID-19 vaccination is a key tool to reduce severe COVID-19^11,12^. New bivalent mRNA vaccines, composed of components of the SARS-CoV-2 ancestral and Omicron BA.5 strain, were made available in the United States in the beginning of September 2022^13^. Bivalent COVID-19 vaccines were recommended by the Advisory Committee on Immunization Practices (ACIP) within the US Centers for Disease Control and Prevention as booster doses to potentially better target the Omicron variant and subvariant waves^14^. Observational clinical data on bivalent COVID-19 vaccines suggest benefit of booster doses of this vaccine to reduce symptomatic infection^6,15^, and additional data supports improved protection against COVID-19 related hospitalizations^16^. As of March 2023, uptake of the new bivalent vaccines in California is low with only 24% of adults and 44% of those over 65 years of age having received a dose^17^.

The oral antiviral drug nirmatrelvir-ritonavir is another key public health tool for minimizing severe COVID-19 outcomes in high-risk patients. In December 2021, nirmatrelvir-ritonavir was given FDA Emergency Use Authorization given evidence that this medication can reduce hospitalization and death among COVID-19 patients with mild/moderate symptoms who are at high-risk for progression to severe COVID-19 within five days of symptom onset^7–9,18^. However, use of nirmatrelvir-ritonavir amongst eligible patients with COVID-19 in the United States has been low, with studies estimating that only 28% of eligible persons were prescribed nirmatrelvir-ritonavir^7,8,10^.

This article reports on the predicted public health impact of increasing uptake of bivalent vaccines and nirmatrelvir-ritonavir in key risk groups on COVID-19 cases, hospitalizations, and deaths. We use the representative case example of California given magnitude of COVID-19 burden and large population size. This study aims to support public health departments to prioritize resources to increase uptake of these interventions in key risk groups to reduce the burden of COVID-19.

## Methods

### Data

We obtained deidentified person-level data on confirmed COVID-19 cases, hospitalizations, and deaths in California from July 23, 2022 to January 23, 2023 from the California Department of Public Health (CDPH). A COVID-19 case was defined as a person whose positive SARS-CoV-2 molecular test was reported to the state. COVID-19 hospitalizations and deaths were defined as a confirmed COVID-19 case who was either hospitalized or died with COVID-19 and reported to the state. CDPH receives reports on hospitalizations and deaths from two independent sources (California COVID-19 Reporting System and healthcare facility-mandated reporting). We used the most inclusive definition of hospitalizations and deaths; we counted a hospitalization or death if either data source indicated that a COVID-19 case led to hospitalization or death. In addition, estimates for hospitalizations utilized a multiplier to account for 75% ascertainment of linkage of cases to hospitalization. We used case episode dates to link the earliest date associated with a SARS-CoV-2 infection for hospitalization and death.

We obtained publicly available vaccine administration data from CDPH (July 23, 2022 - January 23, 2023) to estimate vaccine status, defined as partially vaccinated, fully vaccinated, and boosted. Fully vaccinated referred to those who have received one dose of the Ad26.COV2.S vaccine (Janssen) or two doses of the BNT162b2 mRNA (Pfizer/BioNTech) or mRNA-1273 (Moderna) vaccine. Partially vaccinated referred to those who have received at least one vaccine dose but have not completed a primary series. Boosted referred to those who have completed the primary series and received at least one monovalent vaccine dose. We also estimated age-specific coverage of bivalent booster doses. Vaccine data was demographically stratified by age group (5-11 years, 12-17 years, 18-49 years, 50-64 year, ≥65 years). We excluded vaccination data with missing age information.

### Study Outcomes

The primary study outcomes were COVID-19 cases, hospitalizations, and deaths averted due to one additional vaccine dose or nirmatrelvir-ritonavir treatment.

### Statistical Analysis

We used a modeling approach to predict the number of COVID-19 cases, hospitalizations, and deaths over a future 6-month period to fully capture difference between strategies, and then estimated how many of these outcomes could be averted with additional uptake of bivalent vaccines (or monovalent vaccines for unvaccinated persons) and use of nirmatrelvir-ritonavir during acute illness. We calculated the total averted outcomes (absolute measure), the number needed to treat (relative measure), and proportion of total averted outcomes associated with each intervention strategy.

### Predicting COVID-19 Outcomes

In this model, we predicted the number of cumulative COVID-19 outcomes (cases, hospitalizations, deaths) over the next six months (January 2023 – July 2023) without introduction of any additional vaccination or nirmatrelvir-ritonavir treatment aside from baseline uptake (base case scenario). Models were calibrated to data from July 23, 2022 to January 23, 2023. We used quasi-Poisson regression models to predict the number of weekly COVID-19 outcomes based on age and vaccine status, from which we estimated cumulative COVID-19 outcomes for each age and vaccine status group at the end of the six-month period. We used this parsimonious set of predictors given their strong relation to COVID-19 outcomes^19^ and given our goal was to estimate the cumulative number of COVID-19 outcomes over a time period by relevant risk group (age, vaccination status)^20^. We defined age group as 0-17 years, 18-49 years, 50-64 years, 65-74 years, 75-84 years, ≥85 years, informed by estimations of case fatality rate in each group (see Supplemental Figure A1). We defined vaccine status as unvaccinated, primary series, primary series with one booster dose, primary series with 2 or more booster doses.

Partially vaccinated individuals were classified as fully vaccinated (primary series only) for simplicity as they represented a small fraction (<3%) of total COVID-19 outcomes. We fit separate regression models for each of the three COVID-19 outcomes (cases, hospitalizations, deaths). We accounted for the effect of prior bivalent vaccine coverage during the calibration period, and adjusted the prediction of COVID-19 outcomes based on the expected impact of prior bivalent vaccination (see Supplemental Materials). For model validation, we performed a cross-validation analysis; we used an alternative 6-month calibration period (April 23, 2022 to October 23, 2022) to estimate performance (see Supplemental Table 1).

### Predicting the Impact of Additional Vaccination

We modeled seven vaccination strategies, which simulated administering one additional dose of a COVID-19 vaccine (majority bivalent) to key risk groups, based on age and vaccine status. We modeled the following vaccine strategies, which targeted: 1) everyone, regardless of vaccination status, 2) previously vaccinated individuals, 3) unvaccinated individuals, 4) individuals who have completed the primary series only; 5) 75 years and older (excluding unvaccinated individuals), 6) 65 years and older (excluding unvaccinated individuals); and 7) 50 years and older (excluding unvaccinated individuals). We assumed use of bivalent vaccines for all strategies, except for the primary series which remains as a monovalent dose. These strategies were selected with input from CDPH. While all groups are eligible by guidelines^20^ to receive vaccination, this analysis is intended to provide an estimate of the impact of increasing uptake and coverage in these groups.

Additional analyses were conducted, examining vaccine strategies that stratified older age groups by vaccination status (see Supplement Table 2). For modeling of vaccination, we used available data on vaccine effectiveness of bivalent vaccination by different baseline vaccination statuses against symptomatic infection^6^, hospitalization^16,21^, and death^21^, and extrapolated estimates on vaccine effectiveness from data on monovalent vaccines (assuming this provides a conservative estimate)^21^. We modeled durable vaccine-induced protection over the 6-month simulation period. We did not assume additional benefit beyond 3 booster doses to be conservative. Our assumed vaccine effectiveness estimates for bivalent vaccination are shown in Table 1. We accounted for current baseline coverage of bivalent vaccines (23.7% among eligible population; see Supplemental Materials). We estimated the impact of perfect (100%) coverage of interventions. We calculated total averted outcomes for each vaccine strategy by applying vaccine effectiveness estimates to counts of predicted COVID-19 outcomes by risk groups for different vaccine strategies, subtracting out benefit from recent bivalent vaccination already given. For each vaccine strategy, we calculated the number of vaccine doses necessary for distribution across the California population. We estimated the number needed to treat (NNT) for each vaccine strategy, defined as number of individuals needed to receive an additional vaccine dose to avert one COVID-19 outcome. NNT was calculated as the total vaccine doses administered divided by the total number of outcomes averted.

**Table 1:** Characteristics of COVID-19 cases, hospitalizations, and deaths between July 2022 and January 2023 and model parameters related to vaccination and nirmatrelvir-ritonavir treatment.

|  | Cases<br>(N=1,107,457) | Hospitalizations<br>(N = 45,527) | Deaths<br>(N = 5,650) |
| --- | --- | --- | --- |
| *Descriptive characteristics* |  |  |  |
| **Sex** |  |  |  |
| Female | 595,457 (53.8%) | 23,493 (51.6%) | 2,616 (46.3%) |
| Male | 497,980 (45.0%) | 21,839 (48.0%) | 3,021 (53.5%) |
| Unknown or non-binary | 14,020 (1.2%) | 195 (0.4%) | 13 (0.2%) |
| **Age group (years)** |  |  |  |
| 0-17 years | 133,838 (12.1%) | 1,709 (3.8%) | 27 (0.5%) |
| 18-49 years | 535,518 (48.4%) | 10,320 (22.7%) | 243 (4.3%) |
| 50-64 years | 229,927 (20.8%) | 7,700 (16.9%) | 660 (11.7%) |
| 65-74 years | 106,060 (9.6%) | 8,184 (18.0%) | 1,070 (18.9%) |
| 75-84 years | 65,781 (5.9%) | 9,365 (20.5%) | 1,494 (26.4%) |
| 85+ years | 36,333 (3.2%) | 8,249 (18.1%) | 2,156 (38.2%) |
| **Race/ethnicity** |  |  |  |
| American Indian | 4,132 (0.4%) | 187 (0.4%) | 17 (0.3%) |
| Asian | 120,263 (10.9%) | 4,236 (9.3%) | 806 (14.3%) |
| Black | 52,524 (4.7%) | 3,583 (7.9%) | 391 (6.9%) |
| Latinx | 308,471 (27.9%) | 14,051 (30.9%) | 1,282 (22.7%) |
| Native Hawaiian and<br>other Pacific Islander | 6,641 (0.6%) | 219 (0.5%) | 19 (0.3%) |
| White | 259,414 (23.4%) | 17,697 (38.9%) | 2,857 (50.6%) |
| Multi-Race | 4,918 (0.4%) | 293 (0.6%) | 47 (0.8%) |
| Other | 112,550 (10.2%) | 3,973 (8.7%) | 142 (2.5%) |
| Unknown | 238,544 (21.5%) | 1,288 (2.8%) | 89 (1.6%) |
| **Vaccination status** |  |  |  |
| Unvaccinated | 392,261 (35.4%) | 15,233 (33.5%) | 2,164 (38.3%) |
| Primary Series | 231,386 (20.9%) | 10,624 (23.3%) | 1,172 (20.7%) |
| Boosted (1 dose) | 363,744 (32.9%) | 12,903 (28.3%) | 1,438 (25.5%) |
| Boosted (2+ doses) | 120,066 (10.8%) | 6,767 (14.9%) | 876 (15.5%) |
| *Relative vaccine effectiveness (one additional dose)* |  |  |  |
| **Baseline vaccination status** |  |  |  |
| Unvaccinated (giving one monovalent dose) | 32% (29 – 35)<sup>25</sup> | 29% (21 – 34)<sup>26</sup> | 57% (46 – 69)<sup>26–28</sup> |
| Primary Series (giving one bivalent dose) | 41% (25 – 53)<sup>6</sup> | 57% (41 – 69)<sup>16</sup> | 75% (67 – 83)<sup>29–31</sup> |
| Boosted (1 doses),<br>(giving one bivalent dose) | 26% (1 – 44)<sup>32</sup> | 38% (13 – 56)<sup>16</sup> | 62% (34 – 90)<sup>32–34</sup> |
| Boosted (2 doses),<br>(giving one bivalent dose) | 40% (32 – 47)<sup>6</sup> | 56% (12 – 78)<sup>21</sup> | 63% (27 – 81)<sup>21</sup> |
| *Nirmatrelvir-ritonavir effectiveness* |  |  |  |
| **Vaccination status** |  |  |  |
| Unvaccinated | - | 89% (51 – 100)<sup>35</sup> | 89% (51 – 100)<sup>35</sup> |
| Vaccinated | - | 40% (19 – 56)<sup>8</sup> | 71% (29 – 88)<sup>8</sup> |

See Supplemental Materials for further description of the vaccine effectiveness and nirmatrelvir-ritonavir data. Estimates on vaccine effectiveness are informed by literature from both bivalent and monovalent vaccination.
The category “Other” under the “Race/Ethnicity” section refers to individuals who do not identify with the current available selection of race/ethnicity categories.

### Predicting the Impact of Nirmatrelvir-ritonavir Treatment

We compared three main nirmatrelvir-ritonavir prioritization strategies, which simulate usage of nirmatrelvir-ritonavir during acute COVID-19 in various risk groups. We modeled the following prioritization strategies, which targeted: 1) 50 years and older with co-morbidity or high-risk features, 2) 65 years and older, and 3) 75 years and older. We based medical eligibility for nirmatrelvir-ritonavir as: 1) receipt of a positive SARS-CoV-2 test result, 2) ≤ 5 days since symptom onset or positive test; 3) belonging to a key risk group, such as: ≥ 65 years; ≥50 years and unvaccinated; ≥ 50 with multiple medical co-morbidities; or person with immunocompromising condition. These eligibility criteria were modeled after the FDA’s nirmatrelvir-ritonavir eligibility guidelines^22^. Alternatively, in exploratory analysis, we also investigated: 1) 18 years and older; and 2) 50 years and older— that expanded the eligibility beyond the current guidance (see Supplemental Table 4). In all analyses, we excluded those with contraindications for nirmatrelvir-ritonavir. To account for the true eligible population among each risk group, or the proportion of individuals who meet all the criteria, we created a nirmatrelvir-ritonavir treatment cascade (Figure 2) using published literature estimates for each step. We accounted for current baseline usage of nirmatrelvir-ritonavir (∼22%) and evaluated the effects of increasing prescription (Step D of Figure 2). Additional analyses were conducted, examining nirmatrelvir-ritonavir uptake strategies that stratified older age groups by vaccination status (see Supplemental Table 3).

**Figure 1:**
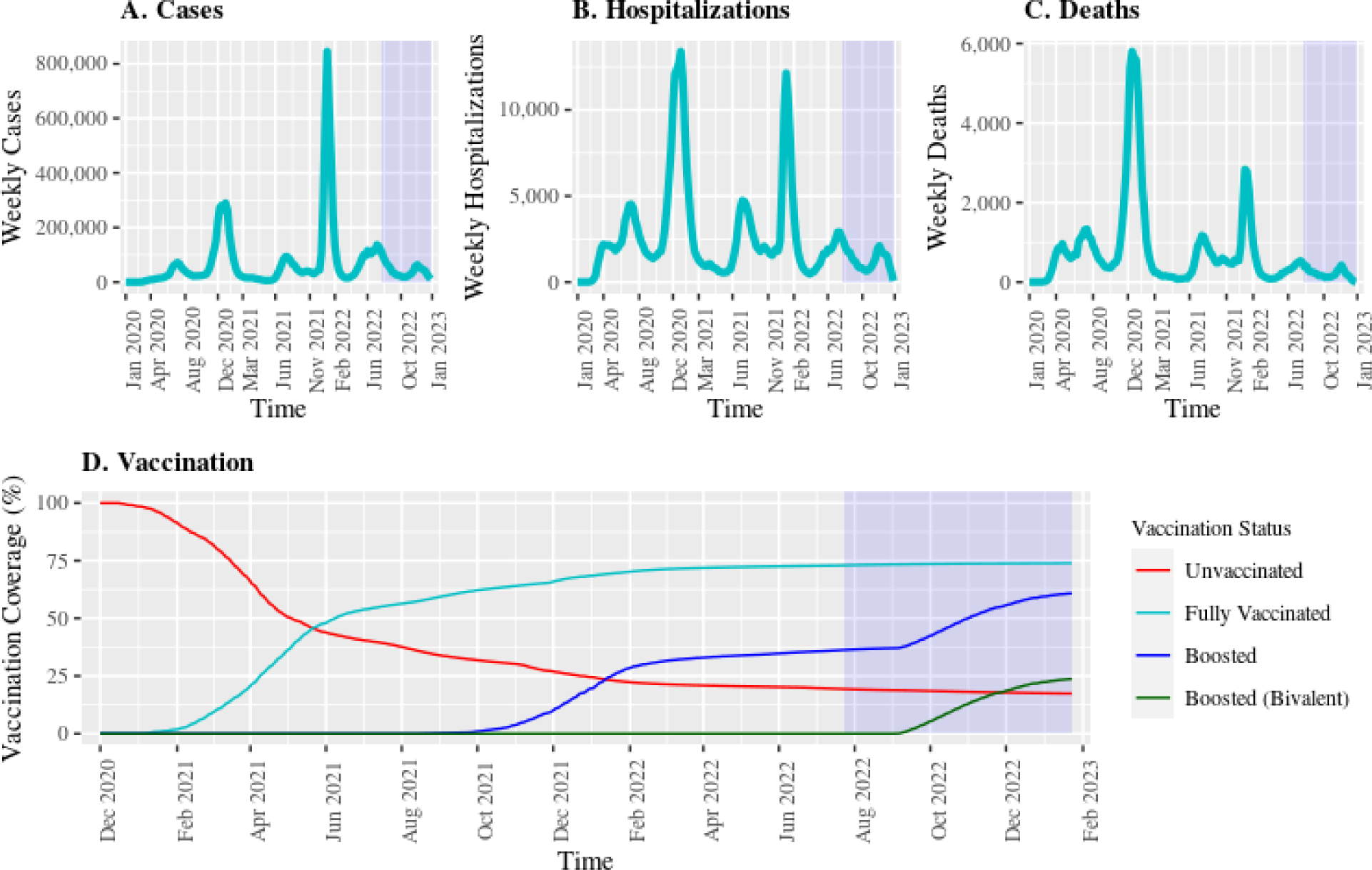
COVID-19 cases, hospitalizations, deaths, and vaccination over time in California. Data on COVID-19 outcomes were obtained from the CPDH for the period of January 1, 2020 to January 23, 2023. This data included weekly absolute COVID-19 cases based on a positive test reported to the state (A), COVID-19 related hospitalizations (B), and COVID-19 related deaths (C). We plotted coverage of different COVID-19 vaccination statuses (D) using publicly available data from December 1, 2020 to January 23, 2023. The boosted coverage is among the booster-eligible population. The model calibration period (July 23, 2022 to January 23, 2023) is shaded in blue.

**Figure 2:**
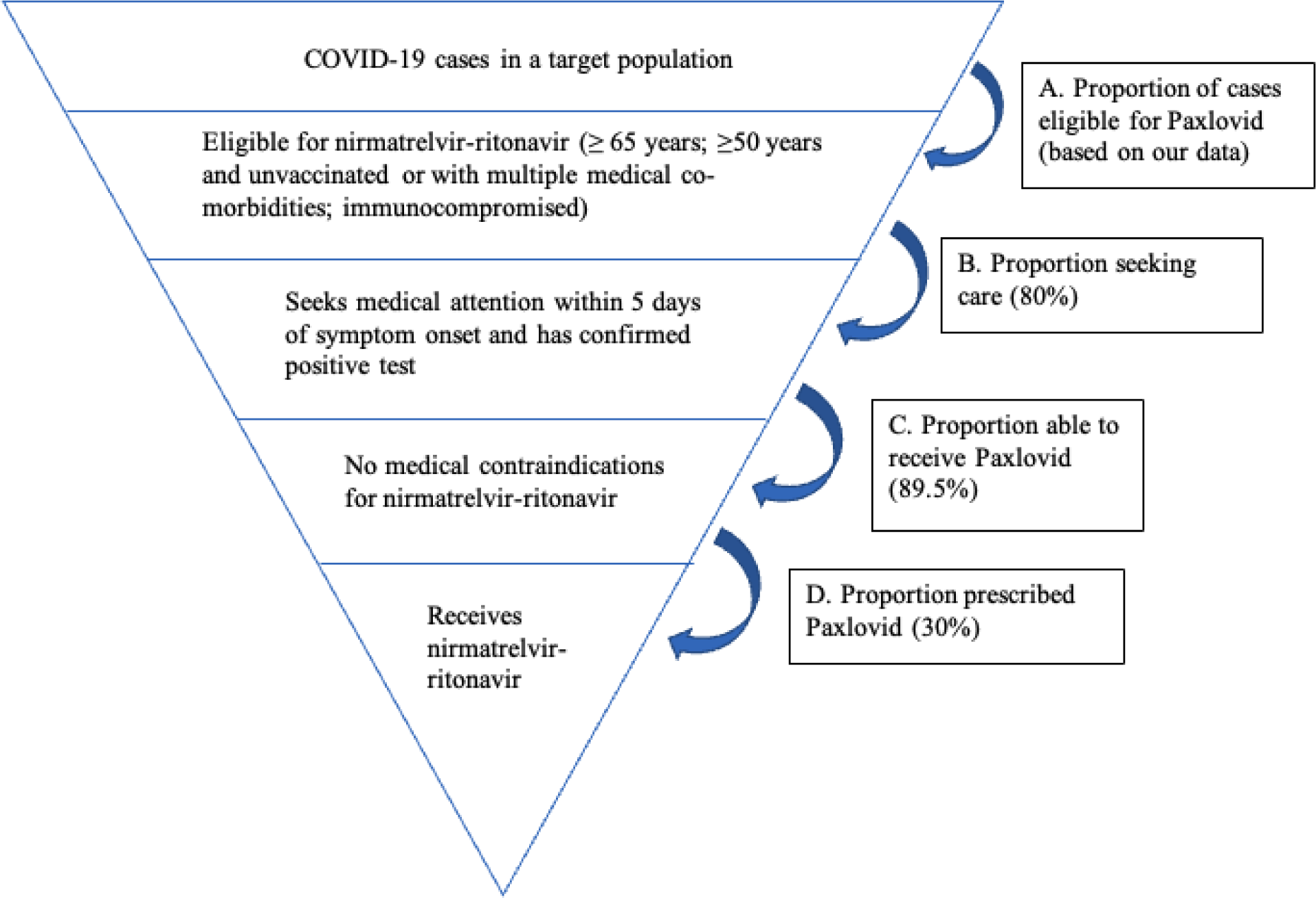
Nirmatrelvir-ritonavir treatment care cascade. We estimated the probability of receiving nirmatrelvir-ritonavir in the population of all COVID-19 cases based on medication eligibility (A), those who seek medical attention (B)^7^, no contraindications (C)^7^, and who are prescribed therapy (D)^7,8^. In the study model, we increased uptake at (D) to simulate higher nirmatrelvir-ritonavir usage.

In this model, we used published data on nirmatrelvir-ritonavir effectiveness to estimate the total number of COVID-19 hospitalizations and deaths averted due to these five different nirmatrelvir-ritonavir prioritization strategies. We extrapolated the effect size of the nirmatrelvir-ritonavir effectiveness for the exploratory analysis in treating persons 18 years and older. Our assumed nirmatrelvir-ritonavir effectiveness estimates for treatment are shown in Table 1. We calculated total averted hospitalizations and deaths for each strategy by applying nirmatrelvir-ritonavir effectiveness estimates to the predicted baseline outcome counts corresponding to the risk groups for different strategies. For each prioritization strategy, we also calculated the number of nirmatrelvir-ritonavir prescriptions necessary for distribution from the COVID-19 case dataset. In addition to estimating total averted outcomes, we also estimated the NNT for each nirmatrelvir-ritonavir prioritization strategy to determine the most treatment-efficient — or optimal — strategy. NNT was calculated as the total prescriptions to persons who otherwise would not have received treatment by strategy divided by the total number of outcomes averted.

### Sensitivity and Uncertainty Analysis

We conducted several sensitivity analyses to evaluate the robustness of our study findings (see Supplemental Materials). We conducted a sensitivity analysis examining waning vaccine-induced immunity against COVID-19 cases (see Supplement Table 5), modeling higher rate of background vaccination (see Supplement Table 6), and different fractions of COVID-19 outcome reporting (see Supplement Table 7). We generated 95% uncertainty intervals for the primary analysis based on uncertainty in COVID-19 outcome and treatment effectiveness (see Supplemental Materials).

### Ethical Approval and Data Sharing

This study was approved by the institutional review board at the University of California, San Francisco. The requirement for informed consent was waived given the analysis used anonymized secondary datasets that were collected as part of public health surveillance and deemed minimal risk. Study reporting followed relevant aspects of Consolidated Health and Economic Evaluation Reporting Standards (CHEERS) guidelines. Data requests can be made to CDPH. Analytic code is available at: github.com/hailey-park/bivalent-vaccines-paxlovid-impacts.

## Results

### Descriptive Data

Over the period from July 23, 2022 to January 23, 2023, there were 1,108,473 confirmed COVID-19 cases reported in California (Figure 1). We excluded 1,016 cases (0.09%) due to missing covariate data; the final sample size was 1,107,457 COVID-19 cases. Among the COVID-19 cases included in this analysis, 45,527 were reported as COVID-19 related hospitalizations (4.1%) and 5,650 were reported as COVID-19 related deaths (0.5%). Over 56% of reported hospitalizations and 83% of reported deaths occurred in those 65 years and older. More information on demographics of COVID-19 cases, hospitalizations, and deaths is shown in Table 1.

As of January 23, 2023, we found that 82% of people in California had received at least 1 dose of a COVID-19 vaccine, 73% of people had completed the primary series, and 60% had received at least one monovalent booster. Uptake of a monovalent booster dose were reported to be highest among adults 18 to 49 years (43%). Coverage of bivalent doses was 23.7% in the overall eligible population, and reported to be highest among those 65 years and older (36% of bivalent doses).

### Model Calibration, Prediction, and Validation

The calibrated regression model predicted a total of 1,151,841 (95% CI: 963,715-1,380,821) COVID-19 cases, 50,426 (95% CI: 43,443-58,575) COVID-19 related hospitalizations, and 6,633 (95% CI: 5,715-7,723) COVID-19 related deaths over a 6-month period, driven by historical data (see Supplemental Figure A2). We performed a cross-validation of the model prediction by changing the calibration periods to an earlier 6-month period (April 23, 2022 – October 23, 2022), and found overall similar relative ranking of risk groups (see Supplemental Table 1).

### Comparison of Vaccine Strategies

We predicted the public health impact of additional COVID-19 vaccine doses and the number needed to avert one COVID-19 case, hospitalization, and death in different epidemiologic groups defined by age and vaccine status (see Table 2). For averting COVID-19 cases, we found the optimal strategy was targeting the unvaccinated group with an additional vaccine dose which averted 125,524 cases (95% UI: 113,288-136,562; NNT: 55), corresponding to 10.9% of total cases. This was followed by targeting everyone which averted 326,111 (95% UI: 249,538-393,601; NNT: 100), and targeting the 75 years and older group which averted 14,318 cases (95% UI: 8,837-18,870; NNT: 106).

**Table 2:** Public health impact and number needed to treat to avert COVID-19 cases, hospitalizations, and deaths with bivalent COVID-19 vaccine strategies.

|  | COVID-19 cases |  |  | COVID-19 hospitalizations |  |  | COVID-19 deaths |  |  |
| --- | --- | --- | --- | --- | --- | --- | --- | --- | --- |
|  | NNT | Total Averted | % Averted | NNT | Total averted | % Averted | NNT | Total averted | % Averted |
| **Everyone** |  |  |  |  |  |  |  |  |  |
| Strategy 1 | 100 | 326,111 | 28.3% | 2,025 | 15,995 | 31.7% | 10,854 | 2,983 | 45.0% |
| *Everyone<sup>a</sup>* | (83 – 130) | (249,538 – 393,601) | (21.7 – 34.2) | (1,707 – 2,881) | (11,241 – 18,975) | (22.3 – 37.6) | (8,304 – 14,560) | (2,224 – 3,899) | (33.5 – 58.8) |
| **Vaccine group-based strategies** |  |  |  |  |  |  |  |  |  |
| Strategy 2 | 128 | 200,588 | 17.4% | 2,212 | 11,561 | 22.9% | 14,619 | 1,749 | 26.4% |
| *Previously vaccinated* | (97 – 205) | (125,016 – 264,472) | (10.9 – 23.0) | (1,749 – 3,615) | (7,074 – 14,621) | (14.0 – 29.0) | (10,368 – 22,180) | (1,153 – 2,466) | (17.4 – 37.2) |
| Strategy 3 | 55 | 125,524 | 10.9% | 1,537 | 4,433 | 8.8% | 5,523 | 1,233 | 18.6% |
| *Unvaccinated<sup>a</sup>* | (50 – 61) | (113,288 – 136,562) | (9.8 – 11.9) | (1,289 – 2,076) | (3,281 – 5,283) | (6.5 – 10.5) | (4,321 – 7,316) | (931 – 1,576) | (14.0 – 23.8) |
| Strategy 4 | 137 | 85,429 | 7.4% | 2,197 | 5,308 | 10.5% | 14,742 | 791 | 11.9% |
| *Primary Series Only* | (105 – 214) | (54,515 – 111,943) | (4.7 – 9.7) | (1,785 – 3,005) | (3,881 – 6,533) | (7.7 – 13.0) | (11,278 – 19,598) | (595 – 1,034) | (9.0 – 15.6) |
| **Age group-based strategies** |  |  |  |  |  |  |  |  |  |
| Strategy 5 | 106 | 14,318 | 1.2% | 387 | 3,920 | 7.8% | 1,410 | 1,074 | 16.2% |
| *75+ years* | (81 – 172) | (8,837 – 18,870) | (0.8 – 1.6) | (311 – 608) | (2,491 – 4,882) | (4.9 – 9.7) | (1,118 – 1,957) | (774 – 1,355) | (11.7 – 20.4) |
| Strategy 6 | 130 | 29,100 | 2.5% | 661 | 5,707 | 11.3% | 2,714 | 1,390 | 21.0% |
| *65+ years* | (99 – 211) | (17,951 – 38,341) | (1.6 – 3.3) | (531 – 1,040) | (3,628 – 7,106) | (7.2 – 14.1) | (2,134 – 3,811) | (990 – 1,768) | (14.9 – 26.7) |
| Strategy 7 | 130 | 68,747 | 6.0% | 1,144 | 7,778 | 15.4% | 5,458 | 1,630 | 24.6% |
| *50+ years* | (98 – 211) | (42,169 – 91,094) | (3.7 – 7.9) | (912 – 1,832) | (4,858 – 9,763) | (9.6 – 19.4) | (4,192 – 7,845) | (1,134 – 2,122) | (17.1 – 32.0) |

<sup>a</sup>For unvaccinated persons, we assumed the vaccine administered was a monovalent dose following current clinical guidance for the primary series.
All analyses compared perfect vaccine uptake to baseline coverage of bivalent vaccines.
All age group-based strategies (Strategies 5-7) excluded the unvaccinated population when targeting vaccines to older age groups.
NNT; number needed to treat.

For averting severe COVID-19 (hospitalization and death), we found the optimal strategy (based on NNT) was targeting the 75 years and older group which averted 3,920 hospitalizations (95% UI: 2,491-4,882; NNT: 387) and 1,074 deaths (95% UI: 774-1,355; NNT: 1,410), corresponding to 7.8% and 16.2% of total hospitalizations and deaths, respectively. The second-best performing strategy was targeting the 65 years and older group, which averted 5,707 hospitalizations (95% UI: 3,628-7,106; NNT: 661) and 1,390 deaths (95% UI: 990-1,768; NNT: 2,714), followed by targeting the 50 years and older group, which averted 7,778 hospitalizations (95% UI: 4,858-9,763; NNT: 1,144) and 1,630 deaths (95% UI: 1,134-2,122; NNT: 5,458).

### Comparison of Nirmatrelvir-ritonavir Strategies

We predicted the public health impact of additional uptake of nirmatrelvir-ritonavir and the number needed to avert one COVID-19 hospitalization and death in different epidemiologic groups defined by age and vaccine status (Table 3). For averting COVID-19 related severe outcomes (hospitalization and death), we found the optimal strategy (based on NNT) was targeting the 75 years and older age group, which averted 5,644 hospitalizations (95% UI: 3,947-6,826; NNT: 11) and 1,669 deaths (95% UI: 1,053-2,038; NNT: 35), and corresponded to 11.2% and 25.2% of total hospitalizations and deaths, respectively. The second-best performing strategy was targeting the 65 years and older group, which averted 8,218 hospitalizations (95% UI: 5,745-9,941; NNT: 15) and 2,160 deaths (95% UI: 1,339-2,661; NNT: 54), corresponding to 16.3% and 32.6% of total hospitalizations and deaths, respectively. The least efficient strategy for averting both severe COVID-19 outcomes was targeting the 50 years and older group, which averted 9,699 hospitalizations (95% UI: 6,882-11,611; NNT: 19) and 2,323 deaths (95% UI: 1,458-2,866; NNT: 79), and corresponded to 19.2% and 35.0% of total hospitalizations and deaths, respectively.

**Table 3:** Public health impact and number needed to treat for nirmatrelvir-ritonavir during COVID-19 infection to avert hospitalizations and deaths.

|  | COVID-19 hospitalizations |  |  | COVID-19 deaths |  |  |
| --- | --- | --- | --- | --- | --- | --- |
|  | NNT | Total averted | % Averted | NNT | Total averted | % Averted |
| **Age group-based strategies** |  |  |  |  |  |  |
| *Based on current eligibility* |  |  |  |  |  |  |
| Strategy 1<br>(50+ years) | 19<br>(16-27) | 9,699<br>(6,882 – 11,611) | 19.2%<br>(13.9 – 22.7) | 79<br>(65 - 126) | 2,323<br>(1,458 – 2,866) | 35.0%<br>(22.5 – 41.8) |
| Strategy 2<br>(65+ years) | 15<br>(12 – 21) | 8,218<br>(5,745 – 9,941) | 16.3%<br>(11.5 – 19.5) | 54<br>(44 – 87) | 2,160<br>(1,339 – 2,661) | 32.6%<br>(20.5 – 39.3) |
| Strategy 3<br>(75+ years) | 11<br>(9 – 15) | 5,644<br>(3,947 – 6,826) | 11.2%<br>(7.9 – 13.4) | 35<br>(29 – 55) | 1,669<br>(1,053 – 2,038) | 25.2%<br>(15.8 – 30.5) |

All analyses compared current uptake (30% in eligible COVID-19 cases without medical contraindications) to 100%.

### Sensitivity Analyses

In our sensitivity analysis incorporating waning vaccine-immunity, we found that the optimal strategies for averting COVID-19 cases were comparable with the primary analysis, with slight worsening in efficiency and absolute impacts (see Supplemental Table 5). When incorporating additional baseline vaccination, we found that the relative ordering of the most optimal strategies was robust, however, with a slight drop in efficiency and absolute impacts (see Supplemental Table 6). Different reporting fractions for COVID-19 outcomes proportionally affected the estimates (see Supplemental Table 7).

## Discussion

In this modeling study, we simulated the public health impact of increasing uptake of bivalent vaccines and nirmatrelvir-ritonavir treatment to avert COVID-19 related cases, hospitalizations, and deaths. We found that targeting those aged 75 years and older with bivalent vaccines and nirmatrelvir-ritonavir treatment during acute illness was the most efficient strategy for minimizing severe COVID-19 related outcomes (hospitalization and death). In general, strategies that prioritized the most high-risk populations (older age groups, unvaccinated individuals) were efficient prioritization strategies, with larger benefits from age-based strategies over vaccine status-based strategies. Higher uptake of bivalent vaccines and nirmatrelvir-ritonavir had similar overall impact, although no strategy entirely averted the burden of severe COVID-19.

We found that for averting severe COVID-19 outcomes using bivalent vaccines, age group-based strategies performed better than vaccine status-based strategies. Targeting vaccines to the unvaccinated population, which may be a challenging strategy, was a high efficiency approach, but its efficiency was still marginally worse than the most inclusive age-based strategy of targeting the 50 years and older group. When comparing these two strategies based on absolute impact, targeting individuals 50 years and older had a larger impact compared to targeting the unvaccinated. The vaccine status-based strategy of targeting those previously vaccinated performed similarly to targeting those 50 years and older based on absolute impact. These results suggest that age, rather than vaccination status, could be emphasized for guidance on prioritization of bivalent vaccines.

We found that nirmatrelvir-ritonavir treatment was projected to be especially impactful for averting COVID-19 related hospitalizations and deaths. When comparing the best-performing strategy of targeting the 75 years and older group between both interventions, the NNT for bivalent vaccines was 387 and 1,410 for hospitalizations and deaths, respectively, whereas the NNT for nirmatrelvir-ritonavir was 11 and 35; this is largely because people taking nirmatrelvir-ritonavir already have confirmed COVID-19 infection whereas vaccination is given to all persons. However, nirmatrelvir-ritonavir under this strategy of targeting the 75 years and older group exhibited higher overall impact when comparing the proportion of total outcomes averted, with bivalent vaccines averting 7.8% and 16.2% of total hospitalizations and deaths, while nirmatrelvir-ritonavir treatment averted 11.2% and 25.2% of total hospitalizations and deaths, respectively. This finding held true when we compared the other age group-based strategies, including targeting the 65 years and older group and the 50 years and older group. Our results suggest that while nirmatrelvir-ritonavir is a high impact intervention (both in terms of efficiency and absolute impacts), both bivalent vaccines and nirmatrelvir-ritonavir treatment are effective interventions. These findings are supported by other nirmatrelvir-ritonavir prioritization studies^23^.We found that expanding eligibility for nirmatrelvir-ritonavir treatment to younger age groups had incremental benefit.

Our study has several limitations. Prospective prediction of COVID-19 outcomes is challenging and our parsimonious model relied on historical data that may not fully capture trends in future COVID-19 outcomes. However, the goal in this study was to compare treatment strategies between risk groups based on historical data; cumulative outcomes over the study period and relative difference between groups was most important to our analysis rather than trends in outcomes. The cross-validation analysis suggests the model may over-predict some COVID-19 outcomes (if COVID-19 outcomes are higher during the calibration period), although the relative ranking of risk groups remained overall consistent. There is limited data on vaccine effectiveness of bivalent COVID-19 vaccines^6,16^, although we made the conservative assumption that these vaccines have at least comparable benefits to booster doses of monovalent vaccines. While benefits of vaccination may wane^24^, the benefits are likely to be sustained over our short study period of 6 months. However, we conducted a sensitivity analysis simulating waning of vaccine-induced protection over time. We also conservatively assumed no additional benefit of 3 or more booster doses due to limited data on relative vaccine effectiveness, although future data can better inform this assumption. Some benefit is likely, which suggests that our results may be underestimating averted outcomes. Our models do not account for prior infection or immunocompromised status due to lack of data, which suggests that our results may overestimate averted outcomes. We applied the same vaccine-induced protection estimates in the immunocompromised population. We assumed complete reporting of all COVID-19 outcomes (except 75% reporting for hospitalization), although there is likely under-ascertainment of COVID-19 cases due to sub-clinical infection and at home rapid antigen tests; our study was most designed to inform strategies to avert severe COVID-19. Finally, the attribution of SARS-CoV-2 infection to reported COVID-19 hospitalizations and deaths remains controversial, although we followed current standard public health classification of these outcomes.

In this study, our findings suggest that prioritizing uptake of bivalent vaccines and nirmatrelvir-ritonavir treatment among the oldest age groups would significantly and efficiently reduce the number of severe COVID-19 infections in California. This study provides evidence on the public health benefit of utilizing both interventions in the United States and highlights potential opportunities for policymakers to improve the promotion and accessibility of these life-saving interventions.

## Data Availability

All data produced in the present study are available upon reasonable request to the authors

https://github.com/hailey-park/bivalent-vaccines-paxlovid-impacts

## Acknowledgements

We thank the California Department of Public Health for sharing the data used in this article and appreciate all the individuals involved in data collection and curation. We specifically appreciate assistance from the CDPH COVID-19 Data Processing and Informatics Section and COVID-19 Modeling Team. We would also like to thank all those involved in the ongoing response to the COVID-19 pandemic in California.

## Disclaimer

This work represents the viewpoints of the authors alone and not necessarily those of the California Department of Public Health, California Health and Human Services Agency, or National Institutes of Health.

## Authorship contribution

Ms. Hailey Park and Dr. Nathan Lo had full access to all the data in the study and take responsibility for the integrity of the data and the accuracy of the data analysis.

Study concept and design: RS, NCL

Statistical analysis: HJP, NCL

Acquisition, analysis, or interpretation of data: All authors

First draft of the manuscript: HJP, NCL

Critical revision of the manuscript: All authors

Contributed intellectual material and approved final draft: All authors

## Previous presentations

None.

