## Appendix for "Predicting the public health impact of bivalent vaccines and nirmatrelvir-ritonavir against COVID-19"

**eAppendix**. Supplemental Methods

**eTable 1**. Cross-validation analysis of primary model to estimate COVID-19 outcomes and relative ranking of risk groups using alternative 6-month calibration period (April 23, 2022 – October 23, 2022) and 3-month validation period (October 23, 2022 – January 23, 2023).

**eTable 2.** Public health impact and number needed to treat to avert COVID-19 cases, hospitalizations, and deaths with bivalent COVID-19 vaccine strategies in risk groups stratified by age and vaccination status.

**eTable 3.** Public health impact and number needed to treat for nirmatrelvir-ritonavir during COVID-19 infection to avert hospitalizations and deaths with stratification of key risk groups by age and vaccination status.

**eTable 4.** Additional strategies for nirmatrelvir-ritonavir treatment with expanded eligibility beyond the current guidance to avert hospitalizations and deaths.

**eTable 5.** Sensitivity analysis of waning vaccine-induced immunity on the public health impact and number needed to treat to avert COVID-19 cases with bivalent COVID-19 vaccine strategies.

**eTable 6.** Sensitivity analysis of status quo vaccination on the public health impact and number needed to treat to avert COVID-19 cases, hospitalizations, and deaths with bivalent COVID-19 vaccine strategies.

**eTable 7.** Sensitivity analysis of case ascertainment on the public health impact and number needed to treat to avert COVID-19 cases with bivalent COVID-19 vaccine strategies.

**eReferences**.

**Contents**

Technical appendix..........................................................................................................................3

Supplemental tables and figures ...…............................................................................................15

**Technical Appendix**

In this appendix, we provide further methodologic detail on the model structure and statistical analysis in this study.

**Additional methodology: Modeling COVID-19 outcomes**

We used a quasi-Poisson regression model to predict the number of weekly COVID-19 outcomes, using age group and vaccination status as our key predictors. We defined age group as 0-17 years, 18-49 years, 50-64 years, 65-74 years, 75-84 years, and ≥85 years, informed by estimates of case fatality rate and case hospitalization rate in each age group (see Figure A1).

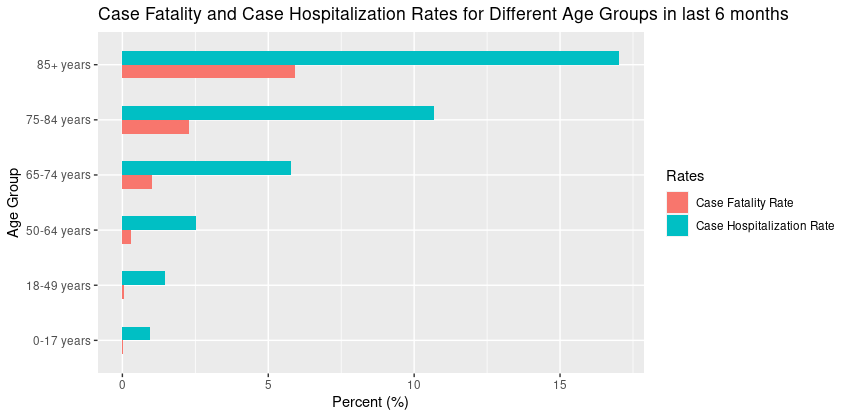

**Figure A1: Case fatality rate and case hospitalization rate for COVID-19 by age group.** Analysis was computed over the time period of July 23, 2022 through January 23, 2023.

*Model Calibration and Prediction*

We fit separate regression models to each of the three COVID-19 outcomes in the study (case, hospitalization, and death). We trained the model on six months of COVID-19 outcomes data in California (July 23, 2022 – January 23, 2023). The calibration plots of the cumulative outcome count for each regression model are shown in Figure A2.

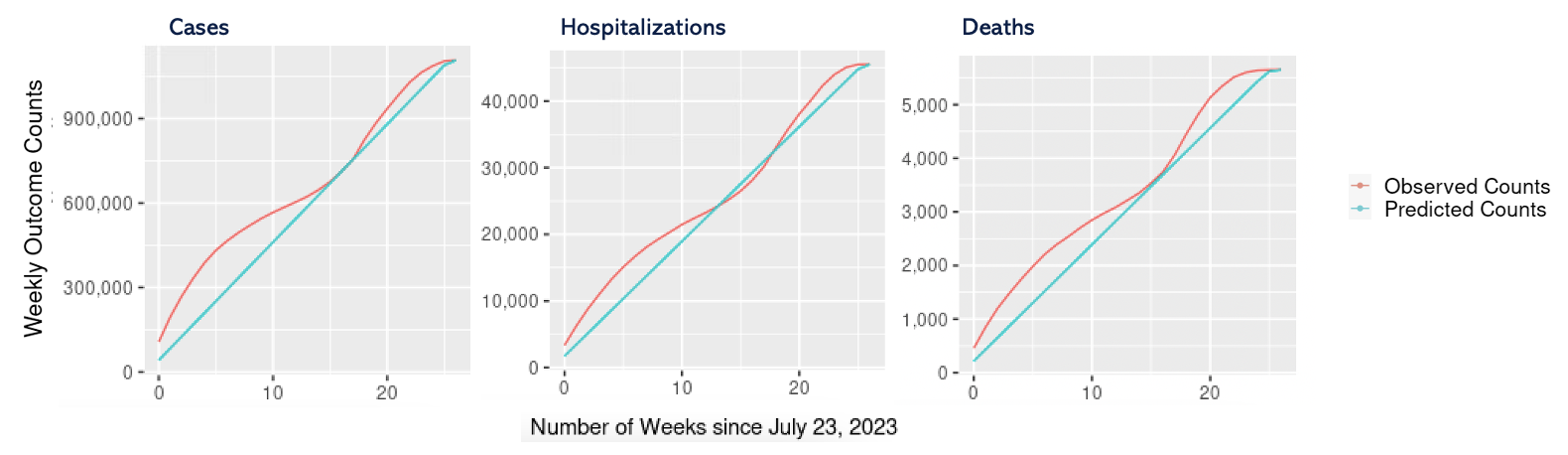

**Figure A2: Model calibration of cumulative COVID-19 outcomes from July 23, 2022 to January 23, 2023.** We provide plots for COVID-19 cases (left), hospitalizations (middle), and deaths (right), each with a different y-axis scale.

We predicted the number of COVID-19 outcomes (cases, hospitalizations, deaths) over the next six months (January 23, 2023 – July 23, 2023) without introduction of any additional vaccination or change in nirmatrelvir-ritonavir treatment uptake (base case scenario), calibrating to data from the preceding 6 months (July 23, 2022 – January 23, 2023). The baseline coverage of COVID-19 bivalent vaccination was 23.7% among those eligible (based on vaccine guidance as of January 23, 2023). We accounted for the effect of prior bivalent coverage on total COVID-19 outcomes observed during the calibration period. We estimated month-specific adjustment factors for each COVID-19 outcome to account for bivalent vaccination, which were calculated using monthly bivalent coverage by age group over the calibration period, uptake of bivalent vaccines by different vaccine groups^1^, prevalence of each vaccine group by age group and month, and relative vaccine effectiveness estimates. These adjustment factors, when applied to the observed outcome data, represent the total COVID-19 outcomes over the calibration period that would have occurred in absence of any bivalent vaccination. The models predicted the number of weekly COVID-19 outcomes in key risk groups by age and vaccine status. Subsequently, we estimated cumulative COVID-19 outcomes for each risk group over a six-month period.

*Cross-validation*

We conducted a cross-validation analysis of our model-predicted COVID-19 outcomes for model validation. The goal was to perform this cross-validation to compare model-predicted and observed COVID-19 outcomes, specifically the relative ranking of COVID-19 outcomes by risk group (defined by age and vaccine status). In this retrospective internal validation analysis, we examined a 9-month period (April 23, 2022 – January 23, 2023). We used 6 months for a training dataset, and 3 months for a test dataset. We calibrated the model to the 6-month period (April 23, 2022 – October 23, 2022); this period captured a time of high COVID-19 incidence and had limited overlap with our original calibration period (July 23, 2022 – January 23, 2023). We validated on the subsequent 3-month period (October 23, 2022 – January 23, 2023). We compared the absolute COVID-19 outcomes and relative rankings by risk group of the model prediction and observed data. The results of this analysis are in Table S1.

**Additional methodology: Modeling interventions against COVID-19**

*Bivalent COVID-19 vaccine effectiveness*

In this model, we used published data on bivalent vaccine effectiveness (and extrapolated monovalent vaccine effectiveness) to estimate the total number of COVID-19 cases, hospitalizations, and deaths averted due to various vaccine strategies. We created composite relative vaccine effectiveness estimates from published literature. For each vaccine effectiveness estimate, if multiple references were used, we fit the study data to a simple random effects meta-analysis model using the R package ‘metafor’ to produce a composite relative vaccine effectiveness estimate. These estimates are shown in Table A1. We used the reported means and 95% confidence intervals of each vaccine effectiveness estimate to fit optimal beta distributions for uncertainty analysis. The mean and the fitted shape parameters of each distribution are shown in Table A2. For the primary series (i.e., vaccine does in previously unvaccinated), we simulated vaccination with the monovalent vaccine following current clinical guidance.

**Table A1: Relative vaccine effectiveness estimates for COVID-19 outcomes.**

|  | Cases | Hospitalizations | Deaths |
| --- | --- | --- | --- |
| Partial Vaccination (Comparator: Unvaccinated) – Monovalent dose | 31.7%  (95% CI: 28.6-34.6)  From the Andrews study (averaged values between Pfizer and Moderna estimates, >4 weeks)^2^  Pfizer: 31.5% (29.9 – 33.1)  Moderna: 31.9% (27.3 – 36.1) | 29%  (95% CI: 21-34)^3^  From the Nyberg study | 57.3%  (95% CI: 45.8 – 68.9)  From the Dagan (72%), Bernal (51%) and Nyberg (42%) study.  42% (-23-72) from the Nyberg study (12+ weeks, deaths)^3^  72% (19-100) from the Dagan study, (14-21 days after 1^st^ dose)^4^  51% (37-62) from the Bernal study (>14 days after 1^st^ dose; converted HR to VE)^5^ |
| 1^st^ Booster: (Comparator:  Primary Series) | 41.0%  (95% CI: 25.0 – 53.0)  From the Link-Gelles study^6^ | 57.0%  (95% CI: 41.0 – 69.0)  From the Tenforde study^7^ | 74.7%  (95% CI: 66.6 – 82.8)  From the Abu-Raddad study on severe outcomes (76.5%), the death estimates from Young-Xu study (74%) and Lin study (74.1%)  76.5% (55.9-87.5) from the Abu-Raddad study^8^  74% (41.1-87.8) from the Young-Xu study^9^  74.1% (62-82.3%) from the Lin study (Supplement Table S5, A)^10^ |
| 2^nd^ Booster (Comparator:  1^st^ Booster) | 25.8%  (95% CI: 1.2 – 44.3)  From the McConeghy study^11^ | 38.0%  (95% CI: 13.0 – 56.0)  From the Tenforde study^7^ | 62.2%  (95% CI: 34.2 – 90.3)  From the death estimates from McConeghy study (89.6%) and Kim study (62.96%) and the severe outcomes estimate from Grewal study (40%).  89.6% (45-100) from the McConeghy study^11^  62.96% (34.2-79.2) from the Kim study^12^  40% (24-52) from the Grewal study^13^ |
| 3^rd^ Booster (Comparator: 2^nd^ Booster) | 40%  (95% CI: 32 – 47)  From the Link-Gelles study^6^ | 56%  (95% CI: 12 - 78)  From the Lin study^14^ | 63%  (95% CI: 27 – 81)  From the Lin study^14^; severe outcomes |

**Table A2: Mean and fitted parameters for the distribution of bivalent COVID-19 vaccine effectiveness by vaccination status.**

| Vaccination Status | | Relative vaccine effectiveness | |
| --- | --- | --- | --- |
| Vaccination Group | Outcome | Mean | Beta distribution (α, β) |
| Unvaccinated^a^ | Cases  Hospitalization  Death | 0.32  0.29  0.57 | Beta (296.3, 630.5)  Beta (51.1, 132.4)  Beta (40.0, 29.6) |
| Primary Series | Cases  Hospitalization  Death | 0.41  0.57  0.75 | Beta (18.0, 27.7)  Beta (26.1, 20.6)  Beta (83.2, 27.3) |
| Boosted (1 dose) | Cases  Hospitalization  Death | 0.26  0.38  0.62 | Beta (3.0, 11.0)  Beta (6.2, 11.6)  Beta (6.3, 3.6) |
| Boosted (2 doses) | Cases  Hospitalization  Death | 0.40  0.56  0.63 | Beta (64.2, 97.9)  Beta (3.6, 3.7)  Beta (6.4, 4.5) |

^a^Assumes monovalent vaccine dose for primary series doses.

*Total vaccine doses for each strategy*

We used publicly available COVID-19 vaccination data and California demographic data to estimate the total number of vaccine doses distributed for each strategy. We assumed California’s total population was 39,240,000^15^. Vaccine administration data was demographically stratified by age group (5-11 years, 12-17 years, 18-49 years, 50-64 year, ≥65 years), and provided cumulative counts by vaccination status (partially vaccinated, fully vaccinated, boosted, and boosted with a bivalent dose). For vaccine strategies that targeted vaccine doses to the oldest age groups (≥75 years, ≥85 years), we extrapolated the total population in these age groups from publicly available metrics on California’s population distribution by age^16,17^.

*Defining key risk groups for nirmatrelvir-ritonavir*

Medical eligibility for nirmatrelvir-ritonavir treatment is often based on clinical evaluation based on being in a key risk group, including individuals: ≥ 65 years; ≥50 years and unvaccinated; ≥ 50 with multiple medical co-morbidities; and those who are immunocompromised. We were able to directly account for age-based and vaccination status using CDPH data on COVID-19 outcome data. We were unable to directly incorporate estimates of COVID-19 outcomes among immunocompromised individuals due to data limitations; however, we used prevalence data from the CDC to account for multiple medical co-morbidities^19^. In addition to belonging to a key risk group, we based medical eligibility for nirmatrelvir-ritonavir as: 1) receipt of a positive SARS-CoV-2 test result, 2) ≤ 5 days since symptom onset or positive test; 3) no contraindications for nirmatrelvir-ritonavir. These eligibility criteria are broadly modeled after the nirmatrelvir-ritonavir eligibility guidelines and common clinical practice.

*Nirmatrelvir-ritonavir effectiveness*

In this model, we used published data on nirmatrelvir-ritonavir effectiveness to estimate the total number of COVID-19 hospitalizations and deaths averted due to various nirmatrelvir-ritonavir prioritization strategies. We used the reported means and 95% confidence intervals of each nirmatrelvir-ritonavir effectiveness estimate to fit optimal beta distributions. The mean and the fitted shape parameters of each distribution are shown in Table A3.

**Table A3: Mean and fitted parameters for the distribution of nirmatrelvir-ritonavir effectiveness against COVID-19 by vaccination group.**

| Vaccination Status | |  | Nirmatrelvir-ritonavir effectiveness | |
| --- | --- | --- | --- | --- |
| Vaccination Group | Outcome | References | Mean | Beta distribution  (α, β) |
| Unvaccinated | Hospitalization  Death | Hammond et al (*NEJM, 2022)*^20^ | 0.89  0.89 | Beta (19.8, 3.4)  Beta (19.8, 3.4) |
| Vaccinated | Hospitalization  Death | Dryden-Peterson et al (*Ann Intern Med, 2022*)^21^ | 0.40  0.71 | Beta (9.5, 15.7)  Beta (4.9, 2.5) |

**Additional methodology: Simulating COVID-19 intervention impact uncertainty**

To quantify uncertainty and generate 95% uncertainty intervals for our model estimate, we used Monte Carlo simulations to account for uncertainty from both regression model predictions of cumulative COVID-19 outcomes and intervention effectiveness (bivalent vaccines, nirmatrelvir-ritonavir treatment). First, we first fit a normal distribution to the mean and 95% confidence interval of the regression model predictions of cumulative COVID-19 outcome for each vaccine and age group. Second, we fit beta distributions to the mean and 95% confidence intervals of vaccine effectiveness estimates (see Bivalent vaccine effectiveness and Table A2) and nirmatrelvir-ritonavir treatment effectiveness (see Nirmatrelvir-ritonavir effectiveness and Table A3). We ran 1000 simulations of intervention effectiveness by randomly sampling values of regression predictions and parameters from fitted distributions. We reported the mean and 95% uncertainty intervals (95% UI) of study outcomes.

**Additional Sensitivity Analyses**

*Stratified analysis by age group and vaccine status*

We conducted an analysis by examining bivalent vaccine and nirmatrelvir-ritonavir uptake strategies that stratified older age groups by vaccination status. For the bivalent vaccine strategies, we stratified the oldest age groups (50+ years, 65+ years, 75+ years) by vaccination status (unvaccinated, primary series only, boosted) to simulate nine total vaccine strategies. For the nirmatrelvir-ritonavir uptake strategies, we stratified the oldest age groups (65+ years, 75+ years) by vaccination status (unvaccinated, primary series only, boosted) to simulate six additional nirmatrelvir-ritonavir strategies. The results of this analysis are in Tables S2-S3 of the Appendix.

*Expanded eligibility for nirmatrelvir-ritonavir*

We simulated additional strategies for expanded eligibility for nirmatrelvir-ritonavir to understand the potential gains in an exploratory analysis, with extrapolation of treatment effectiveness to these new risk groups (outside common clinical practice). We modeled treatment in: 1) 18 years and older, 2) 50 years and older, no eligibility criteria for those 50-64 years — that expanded the eligibility beyond the current guidance. We extrapolated the treatment effectiveness of nirmatrelvir-ritonavir from literature in higher risk populations, which is the assumption underlying this exploratory analysis. The results of this analysis are in Table S4 of the Appendix.

*Waning vaccine-induced immunity*

We conducted a sensitivity analysis modeling the waning of bivalent vaccine-induced immunity against the outcome of COVID-19 case. We only modeled impacts of waning immunity against COVID-19 cases since vaccine-induced protection against severe outcomes (hospitalizations and deaths) over a six month timeframe (length of prediction period) is likely more robust ^22,23^. There is limited clinical data available on the waning effects of bivalent vaccines, so we used available literature on waning of monovalent vaccine effectiveness. We also assumed that a third booster dose has the same waning effectiveness as a second booster dose due to limited clinical data. We created composite relative vaccine effectiveness estimates for bivalent vaccines over time, assuming instantaneous onset of protection and waning immunity at various time points (Table A4). Mean vaccine effectiveness and waning over time of an additional bivalent vaccine dose by baseline vaccination status is shown in Figure A3. We used the reported means and 95% confidence intervals of each vaccine effectiveness estimate to fit optimal beta distributions and simulate study outcomes with Monte Carlo simulations. The mean and the fitted shape parameters of each distribution are shown in Table A5. The results of this analysis are in Table S5 of the Appendix. We assumed no benefit beyond 3 boosters, in part based on limited data.

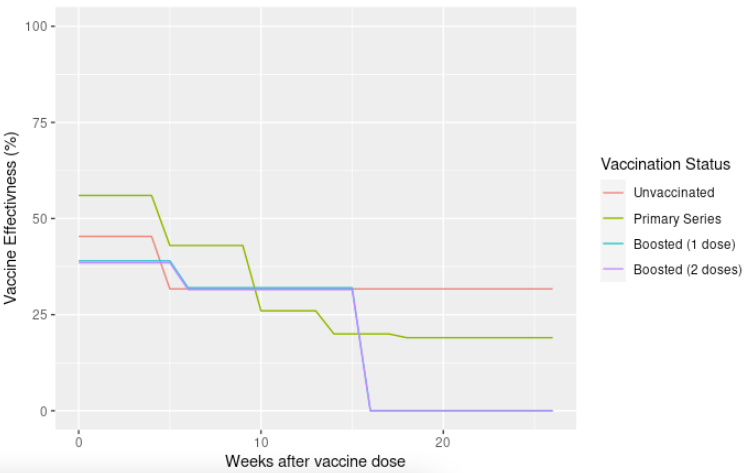

**Figure A3: Relative vaccine effectiveness of COVID-19 bivalent vaccine dose against COVID-19 cases over time by baseline vaccination status.**

**Table A4: Literature on relative vaccine effectiveness of COVID-19 bivalent vaccine dose against COVID-19 cases over time by baseline vaccination status**

|  | Cases |
| --- | --- |
| **Relative Vaccine Effectiveness** |  |
| Partial Vaccination – use of monovalent dose  (Comparator: Unvaccinated) | *Andrews et al.* ^2^  < 4 weeks: 45.35% (41.7 – 48.7)  > 4 weeks: 31.7% (28.6 – 34.6) |
| 1^st^ Booster  (Comparator:  Primary Series) | *Patalon et al.*^24^  1 month: 59.4% (54.9 – 63.5)  2 month: 43.2% (38.2 – 47.8)  3 month: 29.1% (26.1 – 32)  4 month: 18.3% (15.2 – 21.2)  5 month: 16% (12.3 – 19.5)    *Andrews et al.* ^2^  < 4 weeks: 52.2% (47.9 – 56.1)  5-9 weeks: 42.3% (39.3 – 45.2)  > 10 weeks: 22.3% (19.3 – 25.2)    *Composite (Average of both):*  <4 weeks: 55.8% (51.4 - 59.8)  5-9 weeks: 42.8% (38.9 - 46.5)  10-13 weeks: 25.7% (22.7 - 28.6)  14 – 17 weeks: 20.3% (17.3 - 23.2)  >18 weeks: 19.2% (15.8 - 22.4) |
| 2^nd^ Booster  (Comparator: 1^st^ Booster) | *Canetti et al.* ^25^  <5 weeks: 52% (45 – 58)  5-15 weeks: 39% (30 – 45)  >15 weeks: -2% (-27 – 17)    *McConeghy et al.* ^11^  <60 days: 25.8% (1.2 – 44.3)    Composite (average of both):  <5 weeks: 38.9% (23.1 - 51.2)  5-15 weeks: 32.4% (15.6 - 44.6)  >15 weeks: 0% (0 – 17) |
| 3^rd^ Booster  (Comparator: 2^nd^ Booster) | Same as 2^nd^ Booster (Comparator: 1^st^ Booster) waning |

**Table A5: Mean and fitted parameters for the distribution of waning vaccine effectiveness by vaccination status against COVID-19 cases.**

| Vaccination Status | | Relative vaccine effectiveness | |
| --- | --- | --- | --- |
| Vaccination Group | Time since vaccine dose | Mean | Beta distribution (α, β) |
| Unvaccinated | 0-4 weeks  5+ weeks | 0.45  0.32 | Beta (350.8, 424.6)  Beta (291.3, 630.3) |
| Primary Series | 0-4 weeks  5-9 weeks  10-13 weeks  14-17 weeks  18+ weeks | 0.56  0.43  0.26  0.20  0.19 | Beta (298.3, 237.4)  Beta (277.5, 372.1)  Beta (215.3, 624.8)  Beta (143.7, 567.4)  Beta (103.7, 440.2) |
| Boosted (1 dose) | 0-5 weeks  6-15 weeks  16+ weeks | 0.39  0.32  0.00 | Beta (16.7, 28.1)  Beta (11.5, 26.3)  Beta (0.1, 2.0) |
| Boosted (2 doses) | 0-5 weeks  6-15 weeks  16+ weeks | 0.39  0.32  0.00 | Beta (16.7, 28.1)  Beta (11.5, 26.3)  Beta (0.1, 2.0) |

*Higher continued baseline vaccine uptake*

We conducted a sensitivity analysis modeling prospective vaccination, defined as an alternative status quo with increasing COVID-19 vaccine coverage over time. For this analysis, we assumed a 3.3% absolute increase in vaccine coverage each month (20% vaccine coverage by the end of 6 months) in the alternative status quo scenario, instead of assuming complete vaccine coverage at the start of the prediction period. The results of this analysis are in Table S6 of the Appendix.

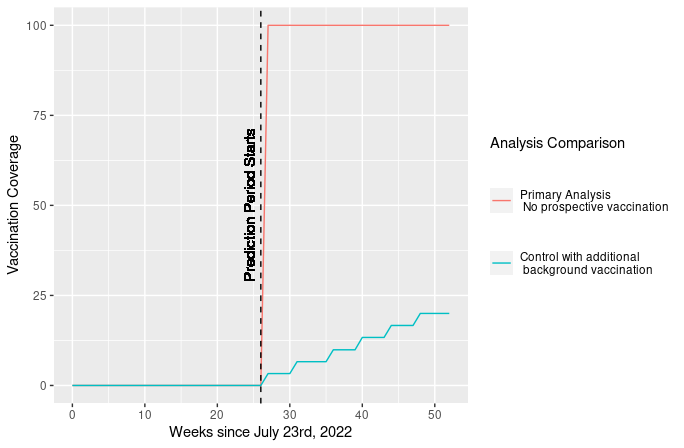

**Figure A4: Comparison of vaccination coverage over time between the primary analysis and continued vaccination sensitivity analysis.**

*Biased case ascertainment*

We conducted a sensitivity analysis to account for low case ascertainment of COVID-19 cases due to sub-clinical infection and at home rapid antigen testing not reported to the state. For this analysis, we assumed COVID-19 case reporting is only at 50%, and applied a fixed two-fold multiplier to case counts. The results of this analysis are in Table S7 of the Appendix.

**Supplemental Tables and Figures**

**Table S1: Cross-validation analysis of primary model to estimate COVID-19 outcomes and relative ranking of risk groups using alternative 6-month calibration period (April 23, 2022 – October 23, 2022) and 3-month validation period (October 23, 2022 – January 23, 2023).**

|  |  | | COVID-19 cases | | |  | COVID-19 hospitalizations | | |  | COVID-19 deaths | | | |
| --- | --- | --- | --- | --- | --- | --- | --- | --- | --- | --- | --- | --- | --- | --- |
|  | Relative ranking  (model) | Relative ranking  (observed) | | Model Prediction  (95% UI) | Observed | Relative ranking  (model) | Relative ranking  (observed) | Model Prediction  (95% UI) | Observed | Relative ranking  (model) | | Relative ranking  (observed) | Model Prediction  (95% UI) | Observed |
| **Everyone** |  |  | |  |  |  |  |  |  |  | |  |  |  |
| Strategy 1  *Everyone* | 1 | 1 | | 925,302  (924,581 – 925,949) | 522,625 | 1 | 1 | 27,736  (27,055 – 28,347) | 25,495 | 1 | | 1 | 3,641  (3,068 – 4,278) | 3,225 |
| **Vaccination group-based strategies** |  |  | |  |  |  |  |  |  |  | |  |  |  |
| Strategy 2  *Previously vaccinated* | 2 | 2 | | 602,728  (602,185 – 603,215) | 356,026 | 3 | 3 | 18,237  (17,725 – 18,696) | 18,570 | 5 | | 4 | 2,236  (1,809 – 2,715) | 2,336 |
| Strategy 3  *Unvaccinated* | 3 | 4 | | 322,574  (322,396 – 322,734) | 166,599 | 5 | 6 | 9,500  (9,331 – 9,651) | 6,925 | 6 | | 6 | 1,405  (1,259 – 1,563) | 886 |
| Strategy 4  *Primary Series Only* | 5 | 6 | | 206,625  (206,446 – 206,786) | 103,796 | 7 | 7 | 6,647  (6,476 – 6,799) | 5,511 | 7 | | 7 | 839  (693 – 998) | 628 |
| **Age group-based strategies** |  |  | |  |  |  |  |  |  |  | |  |  |  |
| Strategy 5  *75+ years* | 7 | 7 | | 55,668  (55,414 – 55,987) | 65,132 | 6 | 5 | 9,445  (9,220 – 9,649) | 11,481 | 4 | | 5 | 2,315  (2,093 – 2,516) | 2,206 |
| Strategy 6  *65+ years* | 6 | 5 | | 127,688  (127,314 – 128,024) | 125,610 | 4 | 4 | 14,079  (13,740 – 14,383) | 16,428 | 3 | | 3 | 2,975  (2,639 – 3,276) | 2,816 |
| Strategy 7  *50+ years* | 4 | 3 | | 306,360  (305,870 – 306,799) | 239,952 | 2 | 2 | 18,835  (18,383 – 19,240) | 20,263 | 2 | | 2 | 3,422  (2,973 – 3,825) | 3,115 |

The cross-validation analysis predicted outcomes over a 3-month period. The observed data refers to the number of COVID-19 outcomes measured over the validation period.

**Table S2: Public health impact and number needed to treat to avert COVID-19 cases, hospitalizations, and deaths with bivalent COVID-19 vaccine strategies in risk groups stratified by age and vaccination status.**

|  | COVID-19 cases | | | COVID-19 hospitalizations | | | COVID-19 deaths | | |
| --- | --- | --- | --- | --- | --- | --- | --- | --- | --- |
|  | NNT  (95% UI) | Total averted  (95% UI) | % Averted  (95% UI) | NNT  (95% UI) | Total averted  (95% UI) | % Averted  (95% UI) | NNT  (95% UI) | Total averted  (95% UI) | % Averted  (95% UI) |
| **Unvaccinated^a^** |  |  |  |  |  |  |  |  |  |
| *Unvaccinated,*  *50+ years* | 24  (22 – 27) | 51,395  (46,358 – 55,946) | 4.5%  (4.0 – 4.9) | 364  (306 – 493) | 3,345  (2,471 – 3,985) | 6.6%  (4.9 – 7.9) | 1,033  (836 – 1,337) | 1,178  (910 – 1,456) | 17.8%  (13.7 – 22.0) |
| *Unvaccinated,*  *65+ years* | 17  (16 – 19) | 25,206  (22,716 – 27,458) | 2.2%  (2.0 – 2.4) | 160  (134 – 217) | 2,628  (1,940 – 3,128) | 5.2%  (3.8 – 6.2) | 402  (327 – 514) | 1,043  (817 – 1,282) | 15.7%  (12.3 – 19.3) |
| *Unvaccinated,*  *75+ years*  **Primary Series** | 14  (13 – 16) | 12,402  (11,167 – 13,522) | 1.1%  (1.0 – 1.2) | 94  (79 – 127) | 1,805  (1,332 – 2,148) | 3.6%  (2.6 – 4.3) | 209  (171 – 265) | 805  (635 – 986) | 12.1%  (9.6 – 14.9) |
| *Primary series only,*  *50+ years* | 100  (77 – 157) | 32,281  (20,590 – 42,278) | 2.8%  (1.8 – 3.7) | 841  (684 – 1,151) | 3,833  (2,800 – 4,713) | 7.6%  (5.6 – 9.3) | 4,306  (3,555 – 5,451) | 748  (591 – 906) | 11.3%  (8.9 – 13.7) |
| *Primary series only,*  *65+ years* | 84  (64 – 131) | 14,811  (9,443 – 19,402) | 1.3%  (0.8 – 1.7) | 423  (345 – 579) | 2,925  (2,138 – 3,595) | 5.8%  (4.2 – 7.1) | 1,897  (1,601 – 2,331) | 652  (531 – 773) | 9.8%  (8.0 – 11.7) |
| *Primary series only,*  *75+ years*  **Boosted** | 69  (52 – 107)) | 7,288  (4,646 – 9,553) | 0.6%  (0.4 – 0.8) | 248  (201 – 338) | 2,009  (1,469 – 2,471) | 4.0%  (2.9 – 4.9) | 985  (842 – 1,186) | 504  (419 – 590) | 7.6%  (6.3 – 8.9) |
| *Boosted,*  *50+ years* | 125  (81 – 319) | 36,466  (14,270 – 56,655) | 3.2%  (1.2 – 4.9) | 1,152  (792 – 3,388) | 3,946  (1,342 – 5,746) | 7.8%  (2.7 – 11.4) | 5,154  (3,563 – 10,310) | 882  (441 – 1,276) | 13.3%  (6.6 – 19.2) |
| *Boosted,*  *65+ years* | 141  (90 – 375) | 14,288  (5,354 – 22,442) | 1.2%  (0.5 – 1.9) | 721  (496 – 2,110) | 2,782  (951 – 4,047) | 5.5%  (1.9 – 8.0) | 2,720  (1,909 – 5,418) | 737  (370 – 1,050) | 11.1%  (5.6 – 15.8) |
| *Boosted,*  *75+ years* | 115  (73 – 306) | 7,030  (2,637 – 11,048) | 0.6%  (0.2 – 1.0) | 421  (290 – 1,233) | 1,911  (653 – 2,779) | 3.8%  (1.3 – 5.5) | 1,412  (997 – 2,785) | 570  (289 – 807) | 8.6%  (4.3 – 12.2) |

^a^Assumes a monovalent vaccine dose for primary series following current clinical guidance.

All analyses compute total averted outcomes based on 100% uptake of vaccination in each group, accounting for baseline vaccine coverage.

NNT; number needed to treat

**Table S3: Public health impact and number needed to treat for nirmatrelvir-ritonavir during COVID-19 infection to avert hospitalizations and deaths with stratification of key risk groups by age and vaccination status.**

|  | COVID-19 hospitalizations | | | COVID-19 deaths | | |
| --- | --- | --- | --- | --- | --- | --- |
|  | NNT  (95% UI) | Total averted  (95% UI) | % Averted  (95% UI) | NNT  (95% UI) | Total averted  (95% UI) | % Averted  (95% UI) |
| *Unvaccinated^a^,*  *65+ years* | 10  (9 – 14) | 4,043  (3,028 – 4,427) | 8.0%  (6.2 – 8.6) | 49  (44 – 65) | 816  (616 – 910) | 12.3%  (9.1 – 13.9) |
| *Unvaccinated^a^,*  *75+ years* | 7  (7 – 10) | 2,776  (2,081 – 3,039) | 5.5%  (4.2 – 5.9) | 31  (28 – 41) | 630  (478 – 695) | 9.5%  (7.1 – 10.8) |
| *Primary series only,*  *65+ years* | 18  (13 – 35) | 1,362  (697 – 1,925) | 2.7%  (1.4 – 3.8) | 59  (43 – 131) | 409  (183 – 561) | 6.2%  (2.9 – 8.1) |
| *Primary series only,*  *75+ years* | 13  (9 – 25) | 935  (478 – 1,322) | 1.9%  (1.0 – 2.6) | 38  (28 – 84) | 316  (142 – 428) | 4.8%  (2.2 – 6.3) |
| *Boosted,*  *65+ years* | 19  (14 – 37) | 2,814  (1,440 – 3,978) | 5.6%  (2.9 – 7.9) | 57  (42 – 126) | 935  (420 – 1,265) | 14.1%  (6.6 – 18.6) |
| *Boosted,*  *75+ years* | 14  (10 – 27) | 1,932  (989 – 2,732) | 3.8%  (2.0 – 5.4) | 36  (27 – 80) | 722  (324 – 970) | 10.9%  (5.1 – 14.5) |

^a^Assumes a monovalent vaccine dose for primary series following current clinical guidance.

All analyses compute total averted outcomes based on comparing baseline treatment uptake (30% in eligible COVID-19 cases) to perfect uptake among those able to be prescribed nirmatrelvir-ritonavir (accounting for care-seeking behaviors, medical contraindications, etc.).

NNT; number needed to treat

**Table S4: Additional strategies for nirmatrelvir-ritonavir treatment with expanded eligibility beyond the current guidance to avert hospitalizations and deaths.**

|  | COVID-19 hospitalizations | | | COVID-19 deaths | | |
| --- | --- | --- | --- | --- | --- | --- |
|  | NNT  (95% UI) | Total averted  (95% UI) | % Averted  (95% UI) | NNT  (95% UI) | Total averted  (95% UI) | % Averted  (95% UI) |
| **Age group-based strategies**  *Not based on current eligibility* |  |  |  |  |  |  |
| Strategy 1  (18+ years) | 39  (32 – 55) | 13,384  (9,353 – 16,192) | 26.5%  (18.7 – 31.7) | 201  (159 – 326) | 2,544  (1,570 – 3,222) | 38.3%  (23.3 – 43.9) |
| Strategy 2  (50+ years) | 23  (19 – 33) | 10,463  (7,309 – 12,660) | 20.7%  (14.6 – 24.8) | 97  (78 – 158) | 2,440  (1,511 – 3,045) | 36.8%  (22.5 – 41.8) |

All analyses compute total averted outcomes based on comparing baseline treatment uptake (30% eligible COVID-19 cases) to perfect uptake among those able to be prescribed nirmatrelvir-ritonavir (accounting for care-seeking behaviors, medical contraindications, etc.).

We extrapolated treatment effectiveness from literature in higher risk groups, and include this as an exploratory analysis as this is outside common clinical practice.

NNT; number needed to treat

**Table S5: Sensitivity analysis of waning vaccine-induced immunity on the public health impact and**

**number needed to treat to avert COVID-19 cases with bivalent COVID-19 vaccine strategies.**

|  | COVID-19 cases | | |
| --- | --- | --- | --- |
|  | NNT  (95% UI) | Total  Averted  (95% UI) | % Averted  (95% UI) |
| **Everyone** |  |  |  |
| Strategy 1  *Everyone* | 116  (105 – 130) | 279,127  (249,758 – 308,387) | 24.2%  (21.7 – 26.8) |
| **Vaccination group-based strategies** |  |  |  |
| Strategy 2  *Previously vaccinated* | 178  (148 – 220) | 144,160  (117,007 – 173,513) | 12.5%  (10.2 – 15.1) |
| Strategy 3  *Unvaccinated^a^* | 51  (48 – 55) | 134,967  (123,841 – 144,109) | 11.7%  (10.8 – 12.5) |
| Strategy 4  *Primary Series Only* | 178  (170 – 188) | 65,596  (62,272 – 68,680) | 5.7%  (5.4 – 6.0) |
| **Age group-based strategies** |  |  |  |
| Strategy 5  *75+ years, excluding unvaccinated* | 143  (122 – 172) | 10,586  (8,829 – 12,485) | 0.9%  (0.8 – 1.1) |
| Strategy 6  *65+ years, excluding unvaccinated* | 176  (149 – 211) | 21,514  (17,948 – 25,359) | 1.9%  (1.6 – 2.2) |
| Strategy 7  *50+ years, excluding unvaccinated* | 177  (149 – 215) | 50,393  (41,469 – 59,933) | 4.4%  (3.6 – 5.2) |

^a^Assumes a monovalent vaccine dose for primary series following current clinical guidance.

This sensitivity analysis models waning of bivalent vaccine-induced immunity against being a COVID-19 case.

All analyses compute total averted outcomes based on 100% uptake of vaccination in each group.

NNT; number needed to treat

**Table S6: Sensitivity analysis with sustained background COVID-19 vaccination on the public health impact and number needed to treat to avert COVID-19 cases, hospitalizations, and deaths with bivalent COVID-19 vaccine strategies.**

|  | COVID-19 cases | | | COVID-19 hospitalizations | | | COVID-19 deaths | | |
| --- | --- | --- | --- | --- | --- | --- | --- | --- | --- |
|  | NNT  (95% UI) | Total  Averted  (95% UI) | % Averted  (95% UI) | NNT  (95% UI) | Total  Averted  (95% UI) | % Averted  (95% UI) | NNT  (95% UI) | Total  Averted  (95% UI) | % Averted  (95% UI) |
| **Everyone** |  |  |  |  |  |  |  |  |  |
| Strategy 1  *Everyone* | 115  (96 – 149) | 283,421  (218,310 – 340,240) | 24.6%  (19.0 – 29.5) | 2,389  (2,024 – 3,355) | 13,556  (9,652 – 16,001) | 26.9%  (19.1 – 31.7) | 12,910  (9,890 – 17,214) | 2,508  (1,881 – 3,274) | 37.8%  (28.4 – 49.4) |
| **Vaccination group-based strategies** |  |  |  |  |  |  |  |  |  |
| Strategy 2  *Previously vaccinated* | 149  (114 – 238) | 172,061  (107,782 – 226,014) | 14.9%  (9.4 – 19.6) | 2,658  (2,110 – 4,294) | 9,622  (5,955 – 12,121) | 19.1%  (11.8 – 24.0) | 18,082  (12,836 – 27,090) | 1,414  (944 – 1,992) | 21.3%  (14.2 – 30.0) |
| Strategy 3  *Unvaccinated^a^* | 62  (57 – 68) | 111,361  (100,508 – 121,150) | 9.7%  (8.7 – 10.5) | 1,731  (1,452 – 2,338) | 3,934  (2,913 – 4,692) | 7.8%  (5.8 – 9.3) | 6,225  (4,868 – 8,275) | 1,094  (823 – 1,399) | 16.5%  (12.4 – 21.1) |
| Strategy 4  *Primary Series Only* | 157  (120 – 246) | 74,416  (47,487 – 97,505) | 6.5%  (4.1 – 8.5) | 2,551  (2,073 – 3,497) | 4,572  (3,336 – 5,627) | 9.1%  (6.6 – 11.2) | 17,275  (13,176 – 22,957) | 675  (508 – 885) | 10.2%  (7.7 – 13.3) |
| **Age group-based strategies** |  |  |  |  |  |  |  |  |  |
| Strategy 5  *75+ years, excluding unvaccinated* | 132  (101 – 213) | 11,499  (7,110 – 15,052) | 1.0%  (0.6 – 1.3) | 484  (393 – 737) | 3,130  (2,054 – 3,854) | 6.2%  (4.1 – 7.6) | 1,771  (1,420 – 2,397) | 855  (632 – 1,066) | 12.9%  (9.5 – 16.1) |
| Strategy 6  *65+ years, excluding unvaccinated* | 162  (124 – 261) | 23,370  (14,457 – 30,602) | 2.0%  (1.3 – 2.7) | 828  (673 – 1,262) | 4,557  (2,993 – 5,612) | 9.0%  (5.9 – 11.1) | 3,411  (2,704 – 4,698) | 1,106  (803 – 1,395) | 16.7%  (12.1 – 21.0) |
| Strategy 7  *50+ years, excluding unvaccinated* | 156  (118 – 255) | 57,066  (34,988 – 75,414) | 5.0%  (3.0 – 6.5) | 1,409  (1,133 – 2,206) | 6,314  (4,035 – 7,854) | 12.5%  (8.0 – 15.6) | 6,791  (5,267 – 9,669) | 1,310  (920 – 1,689) | 19.7%  (13.9 – 25.5) |

^a^Assumes a monovalent vaccine dose for primary series following current clinical guidance.

This sensitivity analysis models a status quo with increasing vaccine uptake over time. For this analysis, we assumed a 3.3% absolute increase in vaccine coverage each month (20% vaccine coverage by the end of 6 months) in the alternative status quo scenario, instead of assuming complete vaccine coverage at the start of the prediction period.

NNT; number needed to treat

**Table S7: Sensitivity analysis on case ascertainment of COVID-19 cases on the public health impact**

**and number needed to treat to avert COVID-19 cases with bivalent COVID-19 vaccine strategies.**

|  | COVID-19 cases | | |
| --- | --- | --- | --- |
|  | NNT  (95% UI) | Total  Averted  (95% UI) | % Averted  (95% UI) |
| **Everyone** |  |  |  |
| Strategy 1  *Everyone* | 50  (42 – 65) | 652,181  (499,251 – 786,688) | 28.3%  (21.7 – 34.2) |
| **Vaccination group-based strategies** |  |  |  |
| Strategy 2  *Previously vaccinated* | 64  (49 – 103) | 401,134  (250,044 – 529,098) | 17.4%  (10.9 – 23.0) |
| Strategy 3  *Unvaccinated^a^* | 28  (25 – 31) | 251,047  (226,682 – 273,009) | 10.9%  (9.8 – 11.9) |
| Strategy 4  *Primary Series Only* | 69  (53 – 107) | 170,835  (109,052 – 223,967) | 7.4%  (4.7 – 9.7) |
| **Age group-based strategies** |  |  |  |
| Strategy 5  *75+ years, excluding unvaccinated* | 53  (41 – 86) | 28,627  (17,655 – 37,696) | 1.2%  (0.8 – 1.6) |
| Strategy 6  *65+ years, excluding unvaccinated* | 65  (50 – 106) | 58,184  (35,876 – 76,630) | 2.5%  (1.6 – 3.3) |
| Strategy 7  *50+ years, excluding unvaccinated* | 65  (49 – 106) | 137,470  (84,337 – 182,219) | 6.0%  (3.7 – 7.9) |

^a^Assumes a monovalent vaccine dose for primary series following current clinical guidance.

This sensitivity analysis models higher case ascertainment (two-fold multiplier).

All analyses compute total averted outcomes based on 100% uptake of vaccination in each group.

NNT; number needed to treat
